# Antimicrobial Susceptibilities of Clinical Bacterial Isolates from Urinary Tract Infections to Fosfomycin and Comparator Antibiotics Determined by Agar Dilution Method and Automated Micro Broth Dilution

**DOI:** 10.1101/2024.07.25.24311029

**Authors:** Jamie L Dombach, Nancy Smith, Teresa Kottiri, Alicia Schiller, Edwin Kamau

## Abstract

Uncomplicated bacteremic urinary tract infections (bUTIs) are common, often caused by *Escherichia coli*, *Klebsiella pneumoniae*, and *Enterococcus faecalis*, with most encounters treated empirically. As rates of antimicrobial resistance increase, available antibiotic treatment options are dwindling. Novel antibiotics approved for treating bUTIs are limited, leading to a resurgence of interest in older antibiotics, including fosfomycin. In this study, clinical urine samples from patients diagnosed with bUTIs from a military hospital on the Eastern Seaboard of the United States were tested for susceptibility to fosfomycin and comparator antibiotics, including levofloxacin, nitrofurantoin, and trimethoprim-sulfamethoxazole (TMS). A total of 1353 nonduplicate bacterial isolates were tested. The majority were Gram-negative, including 605 non-ESBL and 285 ESBL *E. coli* and 84 non-ESBL and 52 ESBL *K. pneumoniae*. Fosfomycin susceptibility rates were similar for non-ESBL and ESBL *E. coli* (95.9% vs 96.1%) and *K. pneumoniae* (38.1% vs 36.5%). Fosfomycin demonstrated high activity against other Enterobacterales and Gram-positive organisms including *Enterobacter faecalis* and *Staphylococcus aureus*. Interestingly, most fosfomycin non-susceptible isolates were susceptible to other first-line bUTI treatment options, and most isolates that were non-susceptible to other first-line bUTI treatment option were susceptible to fosfomycin. ESBL *K. pneumoniae* isolates were the least susceptible to current first-line treatment options. Fosfomycin Etest demonstrated high sensitivity compared to agar dilution, making it a viable method in resource limited areas. Overall, we demonstrated fosfomycin has high activity against common etiologies that cause bUTIs. Further studies investigating the use of fosfomycin in treating non-*E. coli* bUTI pathogens, as single or combination therapy, is warranted.

**IMPORTANCE:** Uncomplicated bUTI are often caused by *Escherichia coli*, *Klebsiella pneumoniae*, and *Enterococcus faecalis*, and treated with antibiotics. As rates of antimicrobial resistance increase, the options available for treatment are diminishing. Limited novel antibiotics have entered the market leading to a resurgence of interest in older antibiotics, including fosfomycin. In this study, we investigated the susceptibility of bUTI clinical isolates to fosfomycin and current treatment. Isolates were susceptible to fosfomycin at similar or higher rates compared to comparator antibiotics, especially for isolates that produce extended-spectrum beta-lactamases. In addition to *E. coli* and *E. faecalis*, organisms that fosfomycin is FDA approved for, fosfomycin had high activity against other Enterobacterales and Gram-positive organisms including *Staphylococcus aureus*. Since most uncomplicated cystitis is treated empirically, fosfomycin is a reasonable treatment option, either single agent or in combination, supporting the need for randomized control trials to approve treatment of other etiologies, and harmonizing breakpoints across agencies.

## INTRODUCTION

Bacteremic urinary tract infections (bUTIs) are common, and can present as asymptomatic or symptomatic uncomplicated, or complicated infections. This distinction is important since it helps guide the choice and duration of antimicrobial treatment (1). Asymptomatic bacteriuria may not warrant treatment, especially in women, and is now recognized as an important contributor to inappropriate antimicrobial use and the emergence of drug resistance (1,2). The most common causes of bUTIs are infections by *Escherichia coli*, *Klebsiella pneumoniae*, and *Enterococcus faecalis*, complicated by the increasing rate of multidrug resistant (MDR) organisms including the extended-spectrum β-lactamase (ESBL) producing strains of *E. coli* and *K. pneumoniae* (3,4). For uncomplicated acute bacterial cystitis, preferred treatments include nitrofurantoin and trimethoprim/sulfamethoxazole (TMS), and more recently fosfomycin, with fluroquinolones and beta-lactam agents used as alternative treatment options (1).

Fosfomycin is a bactericidal antibiotic agent with a broad spectrum activity against Gram-negative and Gram-positive pathogens, including MDR Gram-negative pathogens, such as ESBL-producing organisms and carbapenem-resistant Enterobacterales (CRE) (5,6). Discovered in 1969 (7), the oral formulation (fosfomycin trometamol) was approved by the United States Food and Drug Administration (FDA) in 1996, and is one of the recommended first-line treatment options for uncomplicated bUTIs caused by *E. coli* or *E. faecalis* (1,2).

Due to the limited selection of oral antibiotics to treat bUTIs caused by non-*E. coli* pathogens, especially those that are resistant, fosfomycin usage is increasing (1,6). In a meta-analysis of randomized controlled trials (RCTs) assessing the effectiveness and safety for the treatment of cystitis, fosfomycin trometamol was shown to be as effective with clinical and microbiological cure, relapses, and reinfections as comparator antibiotics including quinolones, β-lactams, aminoglycosides, nitrofurantoin, and sulfonamides (8). Furthermore, observational studies have demonstrated fosfomycin is effective in the treatment of MDR infections (9,10), with oral fosfomycin trometamol having similar clinical and microbiological success with carbapenems in the treatment of ESBL-producing *E. coli* (11). More recently, the FOREST randomized trial showed that fosfomycin trometamol was an effective oral step-down therapy for bUTIs due to MDR *E. coli* compared to other oral drugs (12). With the rates of MDR pathogens increasing worldwide including in the US, the use of fosfomycin for the treatment of bUTIs is on the rise and is likely to increase, warranting additional studies.

Given that bacterial susceptibility profiles can differ regionally, we sought to get a better understanding of the *in vitro* activity of fosfomycin and other first-line antibiotics against bUTI pathogens from the mid-Atlantic region of the United States. In this study we determined the efficacy of fosfomycin, nitrofurantoin, levofloxacin, and trimethoprim/sulfamethoxazole (TMS) against clinical bUTI isolates from a military hospital on the US Eastern Seaboard. Furthermore, we sought to examine the efficacy of fosfomycin against MDR isolates as well as species other than *E. coli* and *E. faecalis*.

## MATERIALS AND METHODS

This study protocol was approved by the appropriate Human subjects regulatory authority. Investigators adhered to the policies for protection of human subjects as prescribed in 45 Code of Federal Regulation 46.

### Bacterial isolates

A total of 1353 nonduplicate bacterial isolates from urine clinical specimens submitted to the clinical laboratory for bUTI testing in our hospital as routine patient standard-of-care were analyzed in this study. Single bacterial colonies obtained from purity plates prepared while setting up automated identification and susceptibility testing on the BD Phoenix system (Becton Dickinson, New Jersey, US) were used for fosfomycin susceptibility testing, which was performed within 24 hours. Fosfomycin susceptibility testing was performed on select isolates at the request of infectious disease clinicians, or randomly selected from the urine bench on the days when fosfomycin testing was performed.

### In vitro susceptibility testing

Susceptibilities of all isolates to fosfomycin (Sigma Chemical Co., St. Louis, MO) was determined by the agar dilution method as described by CLSI, with clinical breakpoints for *E. coli* and *E. faecalis* from the urinary tract set at is ≤64 μg/ml (13,14). For fosfomycin susceptibility testing, plates were prepared using Mueller-Hinton agar (MHA, BBL Microbiology Systems, Cockeysville, MD) supplemented with 25 µg/ml of glucose-6-phosphate and fosfomycin at one of the three concentrations: 64 µg/ml; 128 µg/ml; 256 µg/ml. Bacteria colonies obtained from the purity plates were prepared to 0.5 McFarland and then each isolate spread on four plates; three with fosfomycin (at the three different concentrations) and a control plate that did not contain any antibiotic. Plates were incubated for 16-18 hours at 37°C. The MIC of each antimicrobial agent was defined as the lowest concentration that inhibited visible growth of the organism. Control strains, including *S. aureus* ATCC 29213, *E. coli* ATCC 25922, and *P. aeruginosa* ATCC 27853, were included in each set of tests. Fosfomycin Etest was performed on MHA agar following manufacturer’s recommendations (bioMérieux, Durham, NC).

### Interpretation of susceptibility test results

Interpretive criteria for susceptibility categories by MIC were applied using the CLSI interpretive criteria for urinary tract isolates of *E. coli* and *E. faecalis* for all the isolates as follows: If bacteria grew in the control plate and plate containing 64 µg/ml fosfomycin, but not in 128 and 256 µg/ml fosfomycin, it was considered susceptible; If bacteria grew in the control plate, and in plates containing 64 µg/ml and 128 µg/ml fosfomycin but not 256 µg/ml, it was considered intermediate; if bacteria grew in all the plates including those containing 256 µg/ml fosfomycin, it was considered resistant. Etest endpoints were read by following the manufacturer’s package insert, which permits ≤5 colonies within the ellipse to be ignored.

### Data analysis

MIC data for comparator antibiotics were obtained from the BD Phoenix system and fosfomycin data were obtained from agar dilution and Etest. Isolates that were determined to have intermediate susceptibility were considered non-susceptible for analysis. Descriptive statistics were calculated in Excel and Tableau was used for data representation.

## RESULTS

Of the 1353 bUTI isolates tested, 89.7% (n=1,213) were Gram-negative and 10.3% (n= 140) were Gram-positive. Species with more than 10 bacterial isolates made up 98% (n=1,326) of all organisms tested, with *E. coli*, *K. pneumoniae*, and *E. faecalis* accounting for the majority (84.0%, n=1,137). The remaining species that composed our results were *Pseudomonas aeruginosa*, *Citrobacter spp*, *Proteus mirabilis*, *Enterococcus faecium*, *Enterobacter cloacae*, *Klebsiella aerogenes*, *Klebsiella oxytoca*, and *Staphylococcus aureus* (Figure 1).

**Figure 1.**
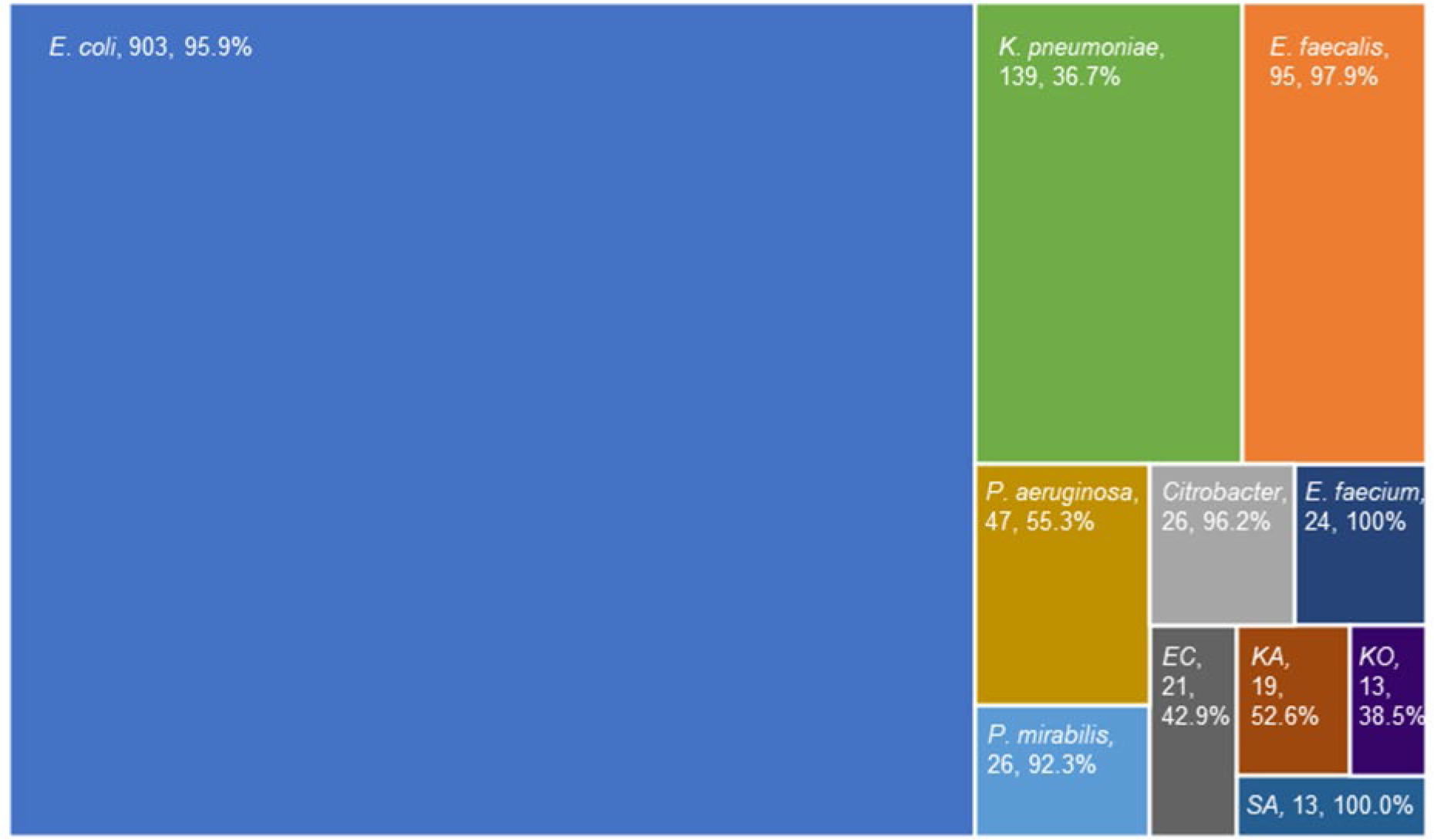
Susceptibility of bacteria isolates from urinary tract infections to fosfomycin. Bacterial isolates (n=1,326) were tested for susceptibility to fosfomycin. *E. coli*, *K. pneumoniae* and *E. faecalis* represented the majority of isolates. Species with more than 10 isolates are displayed (note, different *Citrobacter* species are combined); data shows species, number of isolates tested, and percent susceptible to fosfomycin. EC Enterobacter cloacae; KA Klebsiella aerogenes; SA Staphylococcus aureus; KO Klebsiella oxytoca.

### Antibiotic Susceptibility of Bacterial Isolates

Minimum inhibitory concentration (MIC) values for each antibiotic (fosfomycin, levofloxacin, TMS, and nitrofurantoin) were obtained in 1,326 (of the total 1,353) bacterial isolates (supplementary file 1). Most species exhibited high rates of susceptibility to fosfomycin (92-100%) except for *P. aeruginosa, E. cloacae,* and *Klebsiella spp* which had fosfomycin susceptibilities of less than 56%.

Since *Citrobacter spp* and *P. mirabilis* had high fosfomycin susceptibility rates (96.2% and 92.3%, respectively), and had over 25 isolates, we also evaluated the most common susceptibility patterns in these two species. *Citrobacter spp* overall were more susceptible to the antibiotics tested than *P. mirabilis*. While only 50% of isolates were susceptible to amoxicillin, 92.3% were susceptible to levofloxacin, 84.6% were susceptible to nitrofurantoin, and 88.5% were susceptible to TMS (Supplementary Figure 1). All *Citrobacter spp* isolates were susceptible to at least two antibiotics tested. While only 11.5% of *P. mirabilis* isolates were susceptible to nitrofurantoin, 64.5% were susceptible to levofloxacin, 73.1% were susceptible to TMS, and 96.2% were susceptible to amoxicillin (Supp Figure 1). Like the *Citrobacter spp* isolates, all *P. mirabilis* isolates were susceptible to at least two of the antibiotics tested.

### Antibiotic susceptibility in ESBL vs non-ESBL isolates

Susceptibility profiles for fosfomycin and comparator antibiotics of *E. coli* and *K. pneumoniae* isolates identified as ESBL and non-ESBL strains are represented in Figure 2. There were n=605 non-ESBL and n=285 ESBL *E. coli*, n= 84 non-ESBL and n=52 ESBL *K. pneumoniae* isolates.

**Figure 2.**
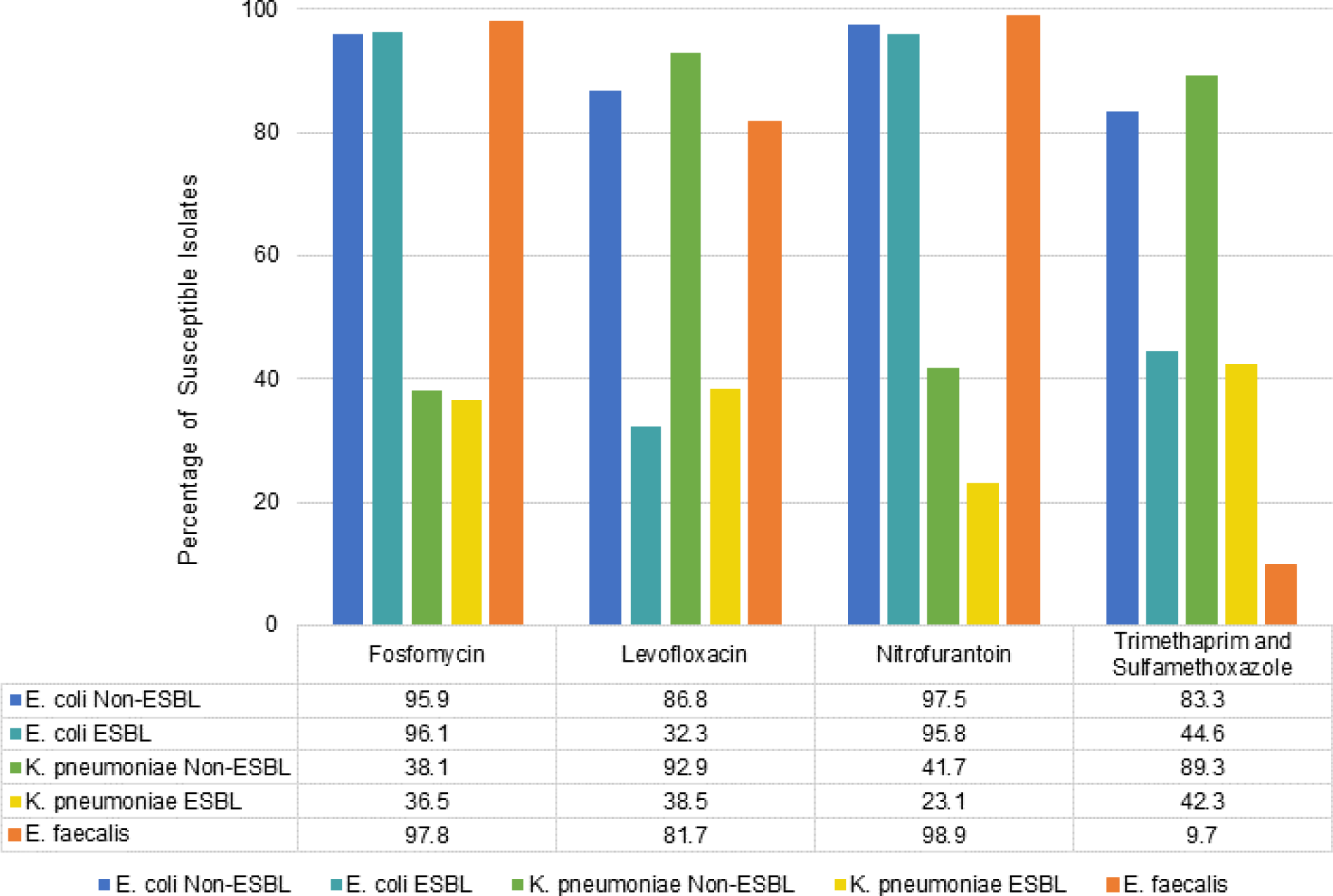
Clinical isolates of *E. coli* and non-ESBL *K. pneumoniae* are susceptible to fosfomycin at similar, or higher rates, than comparator antibiotics. The agar dilution method was used to test whether clinical isolates were susceptible to fosfomycin. Comparator antibiotic susceptibilities were tested using broth dilution. UTI isolates of *E. coli* (n-=890), *K. pneumoniae* (n= 136), and *E. faecalis* (n=93) were tested for susceptibilities to fosfomycin and comparator antibiotics. Isolates of *E. coli* and *K. pneumoniae* were separated by ESBL and non-ESBL to observe antibiotic susceptibilities.

Fosfomycin susceptibility rates were similar for non-ESBL and ESBL *E. coli* (95.9% vs 96.1%) and *K. pneumoniae*, albeit at much lower rates (38.1% vs 36.5%). For the comparator antibiotics, with exception of nitrofurantoin in *E. coli* where susceptibility rates were similar for non-ESBL and ESBL (97.5% vs 95.8%), percentage susceptibility rates for *E. coli* and *K. pneumonia* were about twice or more (≥83.0%) for non-ESBL compared to ESBL (≤44.0%) isolates. Of note, fosfomycin susceptibility profiles appeared to be similar in all species, except for *K. pneumoniae*. The susceptibility profiles of non-ESBL *K. pneumoniae* indicate that levofloxacin and TMS appear to are more effective (Figure 2).

### Susceptibility of multiple-drug resistant (MDR) *E. coli* and *K. pneumoniae* isolates to fosfomycin

We then grouped isolates as MDR based on susceptibility to comparator drugs. When we grouped isolates based on non-susceptibility to levofloxacin and TMS, there were 30 (5.0%) non-ESBL and 121 (42.5%) ESBL *E. coli* MDR isolates, of which all but one non-ESBL and seven ESBL were susceptible to fosfomycin (Figure 3A). For ESBL *K. pneumoniae*, there were 19 (36.5%) MDR isolates, of which 5 (26.3%) were susceptible to fosfomycin. There were only two non-ESBL *K. pneumoniae* MDR isolates, and one was resistant to fosfomycin (Figure 3B). When we grouped isolates based on non-susceptibility to levofloxacin, TMS and nitrofurantoin, there were 5 non-ESBL (0.8%) and 5 (1.8%) ESBL *E. coli* MDR isolates, of which all but one non-ESBL and one ESBL were susceptible to fosfomycin (Figure 3A). For ESBL *K. pneumoniae*, there were 17 (32.7%) MDR isolates, of which 5 (29.4%) were susceptible to fosfomycin. Only one of the non-ESBL *K. pneumoniae* isolates was resistant to all three comparator drugs but it was susceptible to fosfomycin (Figure 3B). Finally, we grouped ESBL *E. coli* and *K. pneumoniae* as MDR based on non-susceptibility to the additional antibiotics tested in the automated AST platform including amoxicillin/clavulanate, aztreonam, cefepime and ceftriaxone. Of the 285 ESBL *E. coli* isolates, 89 were MDR, of which 83 (93.3%) were fosfomycin susceptible. There were 52 ESBL *K. pneumoniae* isolates, 27 were MDR, of which 10 (37.0%) were fosfomycin susceptible.

**Figure 3.**
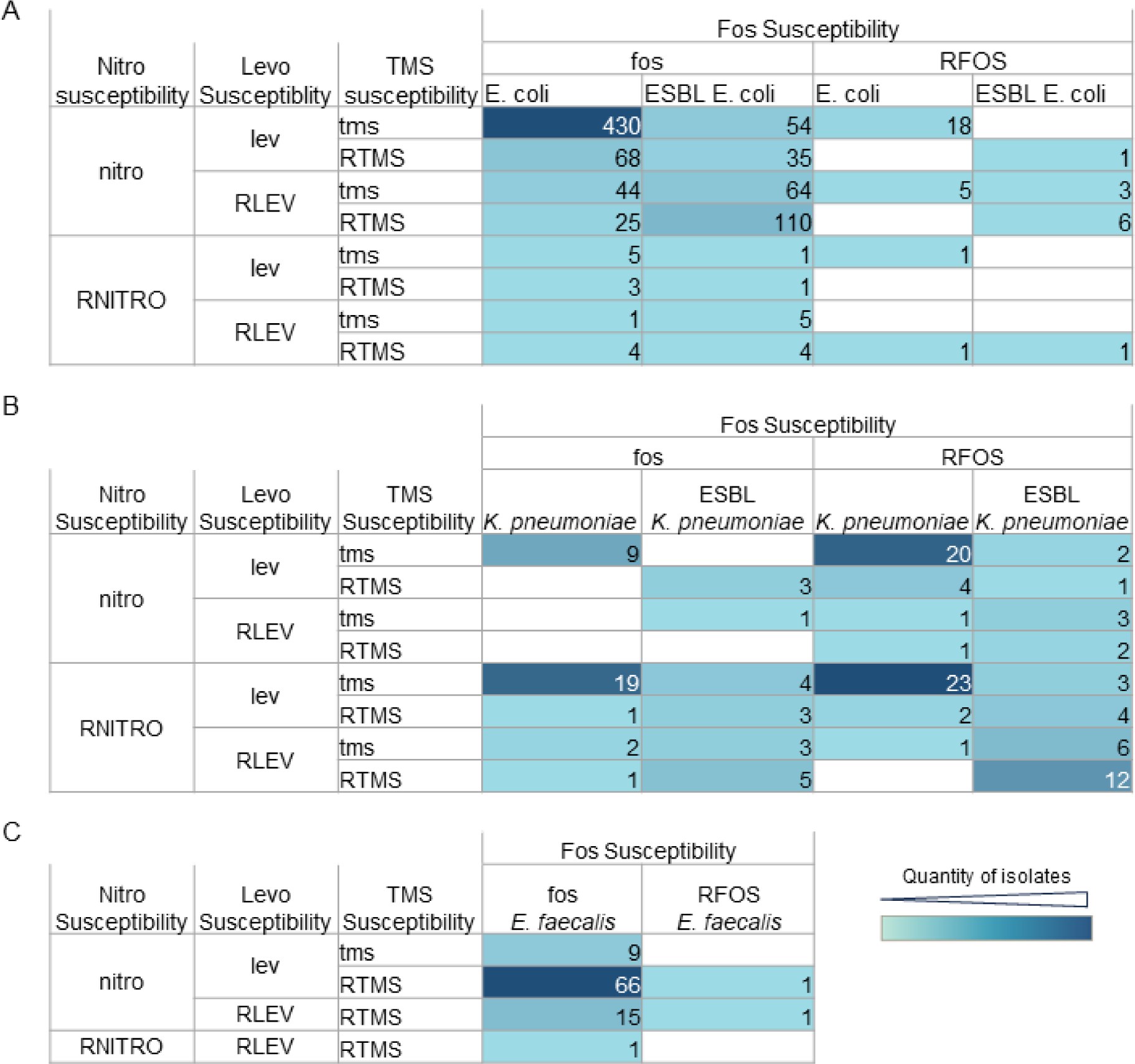
Numerical breakdown of the susceptibility patterns for all isolates tested. Columns depict the number of isolates that are fosfomycin susceptible. Rows further delineate susceptibility to nitrofurantoin, levofloxacin and TMS for *E. coli*, n=890 (A), *K. pneumoniae*, n=136 (B), and *E. faecalis* n=93 (C). fos (fosfomycin susceptible), lev (levofloxacin susceptible), tms (trimethoprim and sulfamethoxazole susceptible), nitro (nitrofurantoin susceptible). RFOS (fosfomycin non-susceptible), RLEV (levofloxacin non-susceptible), RTMS (trimethoprim and sulfamethoxazole non-susceptible), RNITRO (nitrofurantoin non-susceptible).

### Susceptibility of fosfomycin-resistant isolates to comparator drugs

Of the 25 *E. coli* non-ESBL isolates that were fosfomycin resistant, 23 (92.0%) were susceptible to nitrofurantoin and TMS, and of the 15 isolates that were nitrofurantoin resistant, 13 (86.7%) were susceptible to fosfomycin (Figure 3A). Of the 11 *E. coli* ESBL isolates that were fosfomycin resistant, 10 (90.9%) were susceptible to nitrofurantoin, 3 (27.3%) were susceptible to TMS, and only one (9.1%) was susceptible to levofloxacin. Of the 12 nitrofurantoin-resistant ESBL isolates, 11 (91.7%) were susceptible to fosfomycin, and 6 (50.0%) were susceptible to TMS. Of the 52 fosfomycin resistant non-ESBL *K. pneumoniae* isolates, 20 (38.4%) were susceptible to nitrofurantoin, levofloxacin, and TMS, and 43 (82.7%) were susceptible to levofloxacin and TMS. Fosfomycin resistant ESBL *K. pneumoniae* isolates did not display a predominant susceptibility pattern to comparator drugs (Figure 3B).

### Global susceptibility profiles of *E. coli*, *K. pneumoniae* and *E. faecalis*

The global susceptibilities of the isolates to the four antibiotics tested in this study were examined. Isolates were grouped based on the number of antibiotics they were susceptible to. ESBL *K. pneumoniae* displayed the lowest susceptibilities, with 23% of isolates pan-resistant to the four antibiotics tested.

Conversely, non-ESBL *E. coli* exhibited the highest susceptibilities, with 71% of isolates pan-susceptible to all antibiotics, and only one was pan-resistant to all the antibiotics tested. The other three groups of isolates displayed at least 50% susceptibility to at least three antibiotics tested (Figure 4).

**Figure 4.**
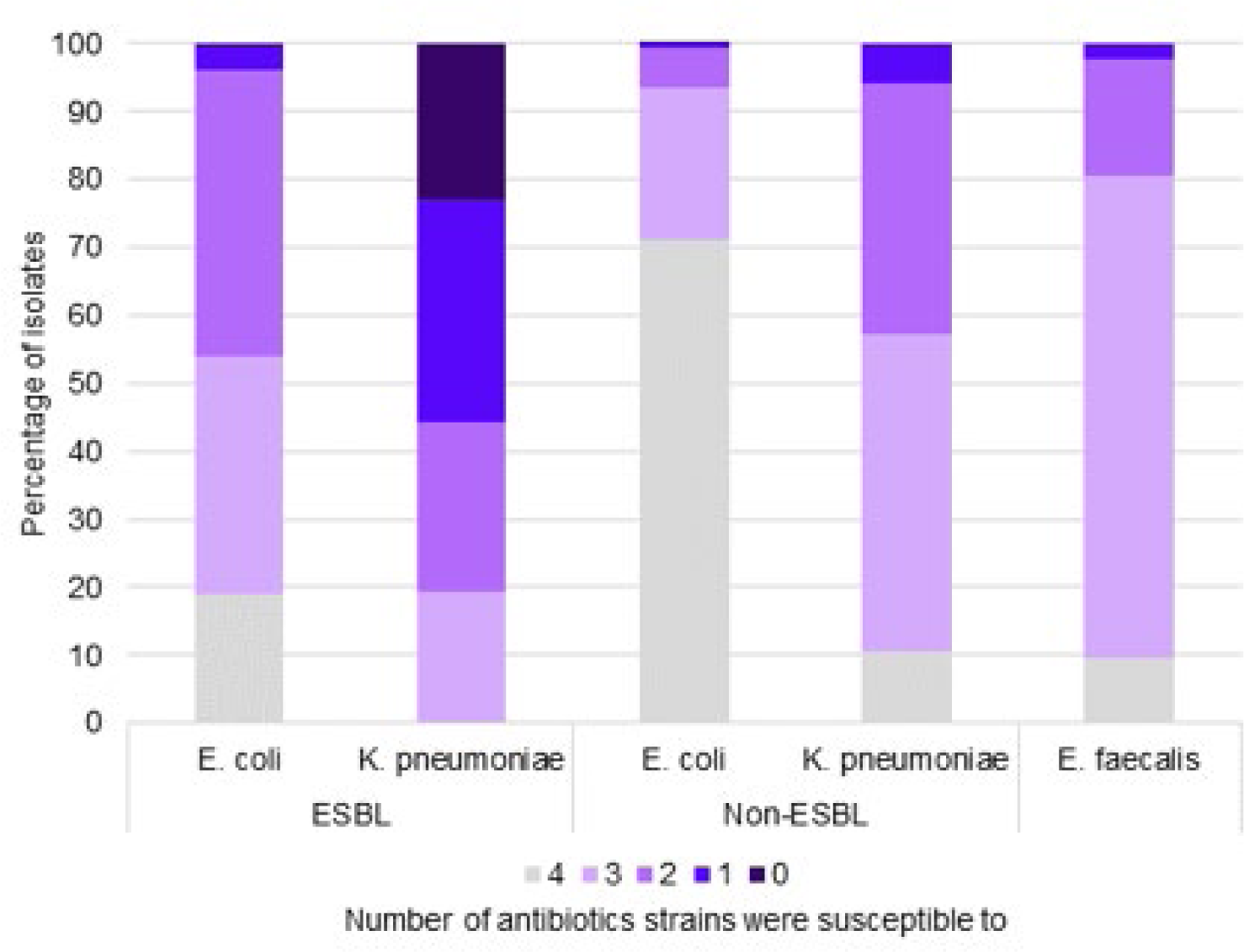
Global susceptibility of isolated strains to the antibiotics tested. Isolates were organized into groups based on the number of antibiotics they were susceptible to. Non-ESBL *E.coli* strains were the most susceptible to the antibiotics tested while ESBL *K. pneumoniae* strains were the least susceptible. Dark purple indicates susceptibility to none of the antibiotics tested and gray indicates strains that were susceptible to four antibiotics.

### Common susceptibility patterns in *E. coli*, *K. pneumoniae* and *E. faecalis*

To understand the most common susceptibility patterns, isolates for each species were grouped by identical antibiotic susceptibilities (Figure 5). The most common susceptibility pattern for non-ESBL *E. coli* isolates was susceptible to fosfomycin, levofloxacin, TMS, and nitrofurantoin, which was present in 430 (71.1%) of the isolates. The most common pattern in ESBL *E. coli* was susceptible to fosfomycin and nitrofurantoin but non-susceptible to levofloxacin and TMS, detected in 110 (38.6%) of the isolates.

**Figure 5.**
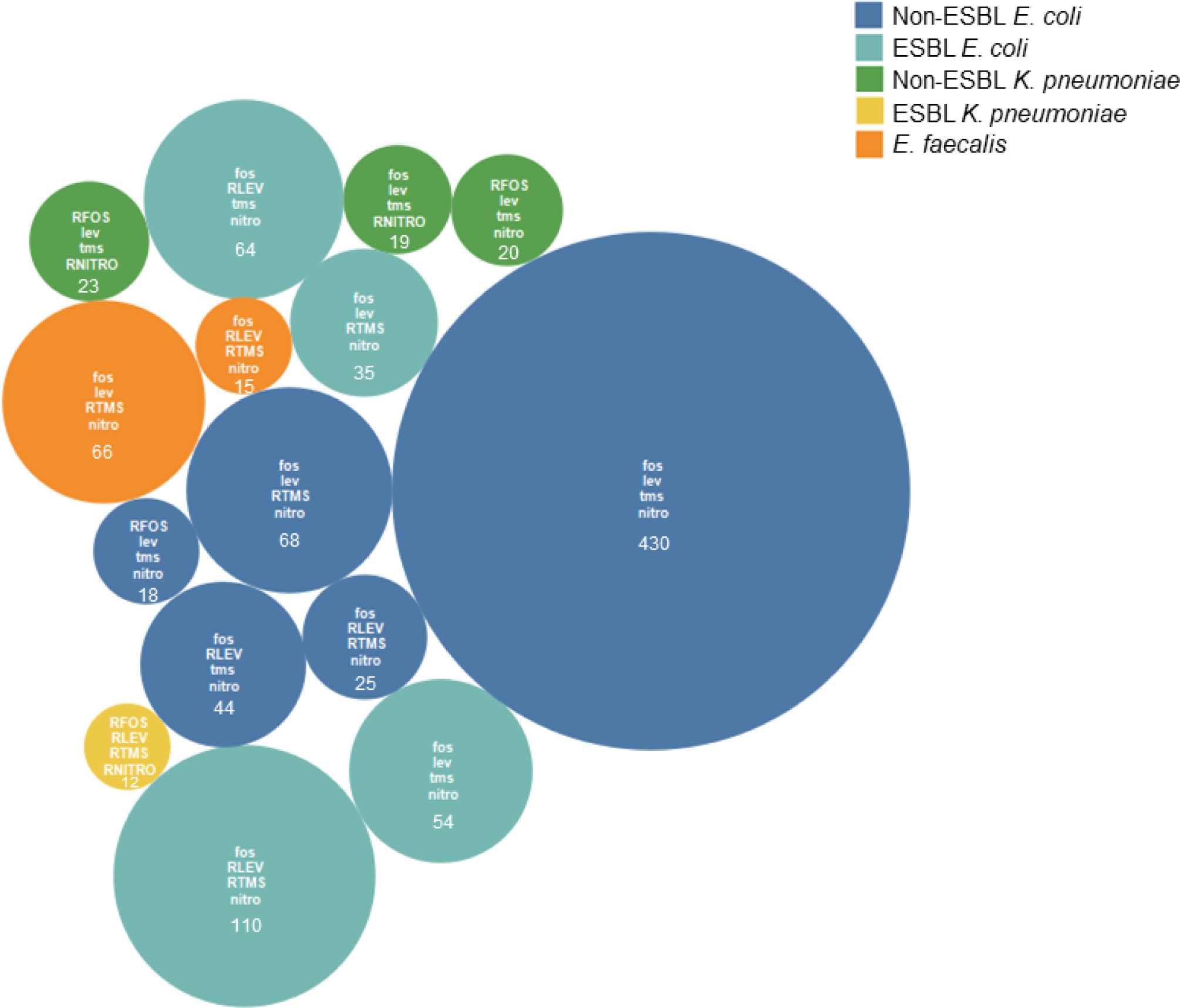
Most common antibiotic susceptibility profiles of tested isolates. Antibiotic susceptibility profiles of all isolates (n=1,119) tested were examined and profiles that had 10 or more isolates are depicted here. The larger the circle the more isolates were in that group. fos (fosfomycin susceptible), lev (levofloxacin susceptible), tms (trimethoprim and sulfamethoxazole susceptible), nitro (nitrofurantoin susceptible). RFOS (fosfomycin non-susceptible), RLEV (levofloxacin non-susceptible), RTMS (trimethoprim and sulfamethoxazole non-susceptible), RNITRO (nitrofurantoin non-susceptible). Number of isolates for each profile included.

While three susceptibility patterns were prominent in non-ESBL *K. pneumoniae,* the most common pattern, with 23 (27.4%) isolates, was susceptible to levofloxacin and TMS but non-susceptible to fosfomycin and nitrofurantoin. Interestingly, the susceptibility pattern that was most common in ESBL *K. pneumoniae* was non-susceptible to all four antibiotics tested. ESBL *K. pneumoniae* exhibited the highest number of susceptibility patterns, 14 out of the possible 16 patterns represented (Figure 3). The majority of *E. faecalis* isolates (66 [71.0%]) were susceptible to fosfomycin, levofloxacin, and nitrofurantoin but non-susceptible to TMS.

### Comparison of agar dilution and Etest outcomes

Given that agar dilution (AD) is time and material-intensive, we tested if the Etest, which may be more practical for many labs performing surveillance, especially in resource-limited areas, could provide accurate results. Fosfomycin susceptibility profiles were established using the Etest in the following bacterial isolates: 477 *E. coli*, of which 34 were ESBLs; 96 *K. pneumoniae*, of which 11 were ESBLs; and 45 *E. faecalis*. There was a dramatic range in MICs (Figure 6), which led to more considerable than expected disparities between the median and average MICs (Table 1). Overall *E. coli* was highly susceptible compared to *K. pneumoniae* for non-ESBLs (99.8% vs. 50.6%) and ESBLs (91.0% vs. 45.5%) and *E. faecalis* had high susceptibility rates (88.9%) (Table 1). Of the 116 isolates tested for fosfomycin susceptibility by both Etest and AD, 116 (100.0%) were identified as susceptible by AD but only 109 (94.0%) were identified as susceptible by Etest. Three misidentified isolates had MICs greater than 256 µg/mL.

**Figure 6.**
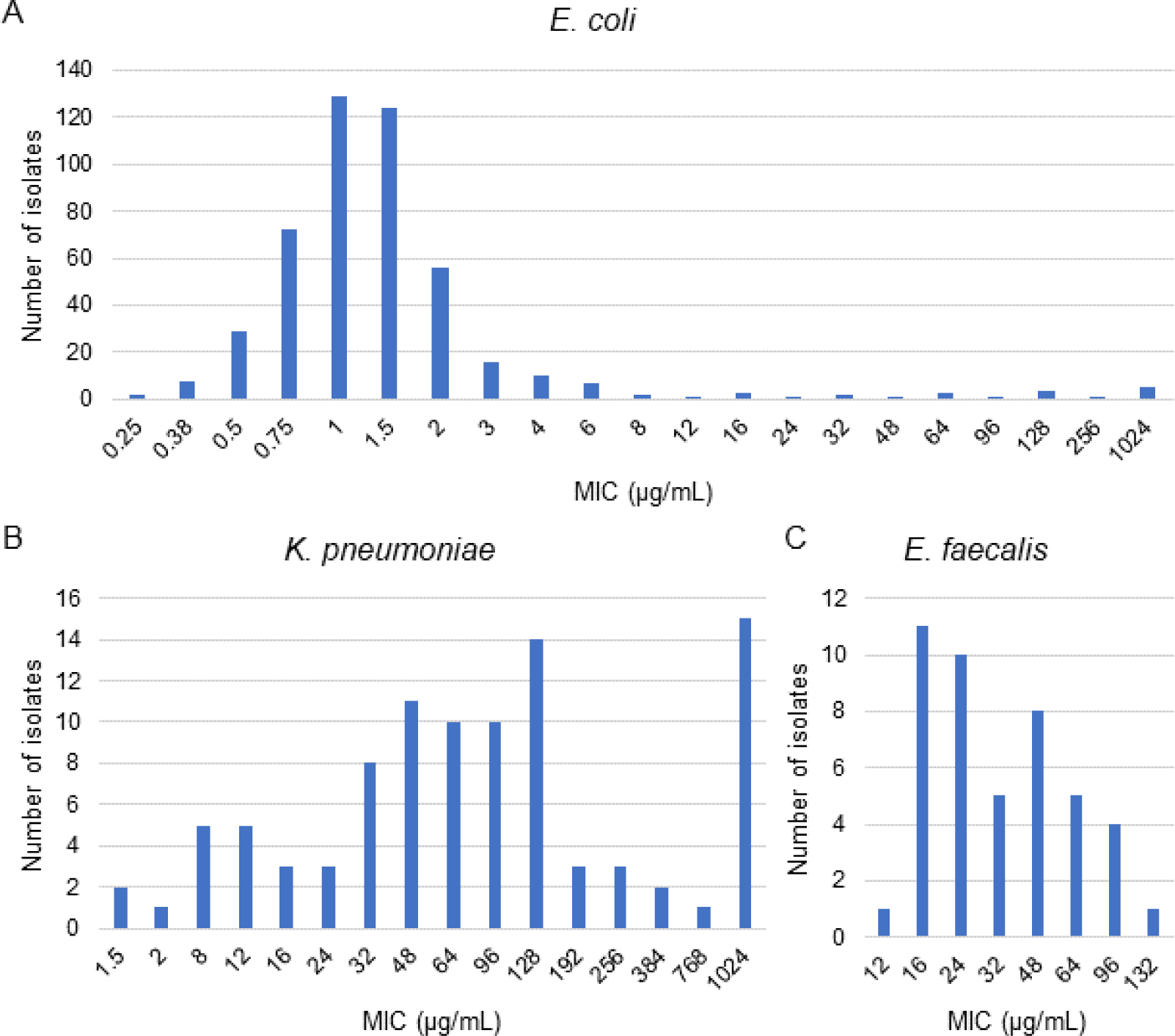
Distribution of fosfomycin MICs. MICs for fosfomycin were determined by E test for (A) *E. coli*, (B) *K. pneumoniae*, and (C) *E. faecalis*.

**Table 1.** Isolate susceptibility to fosfomycin determined by the Etest.

|  | Non-ESBL |  | ESBL |  |  |
| --- | --- | --- | --- | --- | --- |
|  | *E. coli* | *K. pneumoniae* | *E. coli* | *K. pneumoniae* | *E. faecalis* |
| **Median** | 1 | 96 | 1.5 | 64 | 32 |
| **Average** | 15.2 | 255.8 | 8.22 | 81.8 | 40.2 |
| **Standard Deviation** | 109.3 | 372.5 | 24.1 | 57.3 | 28.0 |
| **Percent susceptible** | 99.8 | 50.6 | 91.0 | 45.5 | 88.9 |
| **Percent susceptible (AD)** | 95.9 | 36.4 | 96.4 | 37.3 | 98.0 |

## DISCUSSION

In this study, we evaluated fosfomycin activity against common bUTI organisms collected from a military hospital located in the Eastern Seaboard of the United States using CLSI breakpoints for *E. coli* and *E. faecalis*. In addition to showing high activity against these two organisms, fosfomycin also displayed high activity against other common bUTI organisms, including Enterobacterales (*Citrobacter spp* and *P. mirabilis*) and Gram-positive organisms (*S. aureus* and *E. faecium*). Further, fosfomycin displayed high activity against ESBL and MDR *E. coli* isolates, with similar susceptibility rates as those for non-ESBL isolates. Nitrofurantoin displayed similarly high activity against *E. coli* as fosfomycin. Interestingly, most fosfomycin non-susceptible isolates were susceptible to nitrofurantoin, and most nitrofurantoin non-susceptible isolates were susceptible to fosfomycin. Fosfomycin displayed least activity against *K. pneumoniae*, both non-ESBL and ESBL isolates. Fosfomycin non-susceptible non-ESBL *K. pneumoniae* were highly susceptible to levofloxacin and TMS but not nitrofurantoin. Fosfomycin also displayed low activity against *K. aerogenes*, *E. cloacae* and *P. aeruginosa*. The susceptibility rates in our study corroborated a recent study which showed *P. aeruginosa* with susceptibility rate of 40.4% to fosfomycin (15).

As a primary or step-down bUTI treatment, fosfomycin has been shown to have high clinical, microbiological, and *in vitro* activity against MDR *E. coli*, including those harboring metallo-β-lactamases (12, 16–26). Oral fosfomycin tromethamine is FDA-approved for single-dose treatment of uncomplicated bUTI (23,27). As the predominant bUTI pathogen, most published studies investigating clinical, microbiological, and *in vitro* activity of fosfomycin have primarily focused on *E. coli* (28). There are however studies that have also investigated activity of fosfomycin against other common bUTI species. A study by Mezzatesta *et al*., found in addition to having high activity against ESBL *E. coli*, fosfomycin had high *in vitro* activity against ESBL *P. mirabilis* and methicillin-resistant *S. saprophyticus* (5). Other studies have also demonstrated high fosfomycin *in vitro* activity against Enterobacterales, including *Proteus spp*, *Enterobacter spp*, *Serratia spp*, and *Citrobacter spp* (28,29). These studies corroborate part of our data where we demonstrated high fosfomycin *in vitro* activity against *P. mirabilis* and *Citrobacter spp* but not *Enterobacter*. Further, similarly to our study, studies have demonstrated that fosfomycin retains high *in vitro* activity against bacterial isolates that are non-susceptible to other first-line treatments for bUTI (28,29).

Fosfomycin *in vitro* activity against *K. pneumoniae* seems to vary widely between studies but remains consistently lower than that of most other Enterobacterales (5, 27–30). Interestingly, high clinical and microbiological cures have been attained in patients treated with fosfomycin due to bUTI caused by *K. pneumoniae* (31–33). Although the epidemiology and susceptibility of different organisms may vary based on the country and region, these data strongly suggest fosfomycin is a highly active antibiotic against the most common causes of bUTI and has a high clinical cure rate. This is important since most uncomplicated bUTI are treated empirically (34). Therefore, an antibiotic that provides broad-spectrum activity, including activity against MDR organisms with good clinical outcomes, is a reasonable treatment option. Further, fosfomycin is a highly viable option as a step-down treatment (12,18,20). RCTs will be invaluable in investigating fosfomycin effectiveness in treating other common bUTI pathogens including *K. pneumoniae*, especially in regions with high rates of MDR organisms.

Fosfomycin tromethamine is an antibacterial phosphonic acid that inhibits bacterial cell wall formation by interfering with peptidoglycan synthesis (35). *In vitro* synergy between fosfomycin and other antibiotics has been demonstrated, which may be useful as an alternative treatment option, especially for MDR organisms including *K. pneumoniae*, *P. aeruginosa*, *Acinetobacter baumannii* and methicillin-resistant *S. aureus* (36). The synergistic effect between fosfomycin and β-lactam antibiotics is proposed to arise from the inhibition of cell wall synthesis at separate steps, and ciprofloxacin is due to ciprofloxacin-mediated damage to the outer membrane, which increases the penetration and activity of fosfomycin (35,37–38). In *P. aeruginosa*, *in vitro* studies demonstrated synergy between fosfomycin and a variety of other antibiotics, including aztreonam, cefepime, meropenem, imipenem, ceftazidime, gentamycin, amikacin, ciprofloxacin, and others (37–40). Further, the synergistic activity of fosfomycin, ciprofloxacin, and gentamicin against *E. coli* and *P. aeruginosa* biofilms has been demonstrated (41), making it a reasonable treatment option for patients with catheters. RCTs investigating fosfomycin as a combination therapy have shown higher clinical and microbiological success rates in many different infections, including sepsis/bacteremia and respiratory tract infections (25, 42,43). However, to the best of our knowledge, RCTs investigating fosfomycin as a combination therapy in treating UTIs have not been conducted (42). In our study, with the exception of one isolate, all fosfomycin-resistant *E. coli* were susceptible to either all or at least one of the comparator UTIs first-line or alternate drugs. Clinical studies have compared the efficacy of fosfomycin to comparator UTI first-line drugs and have shown it to be a valuable alternative option (24, 42, 44–47). In a study investigating treatment patterns and clinical outcomes among females with uncomplicated bUTI in Germany that were empirically treated, the incidence of recurrent infection in the 6-month post-index period was higher in patients with *E. coli* that were resistant to one of the first-line treatments (48). With antimicrobial susceptibility rates and epidemiology of bUTI pathogens different and rapidly changing from one region to another, robust research with well-designed double-blinded RCTs investigating the use of fosfomycin as a combination therapy for the treatment of bUTIs, especially in patients with recurrent infections, is warranted.

Similar to the European Committee on Antimicrobial Susceptibility Testing (EUCAST), the Clinical and Laboratory Standards Institute (CLSI) recognizes AD as the reference method for fosfomycin susceptibility testing for *E. coli* and *E. faecalis* (13,49,50). AD is a labor- and time-intensive methodology, which has led to many laboratories using either disk diffusion, which is only approved for *E. coli* from urine samples by CLSI (50), or Etest, which is only approved by EUCAST (49) but not CLSI. This is further complicated by differences in breakpoints between CLSI and EUCAST, which also varies by specimen types (51). Etests, based on a combination of dilution and diffusion testing, offer valuable solutions since they provide MICs. However, there are concerns about the accuracy of Etest results, which can also be challenging to interpret due to micro- and macro-colonies (51, 52). A recent new formulation of the fosfomycin Etest showed high accuracy, which is vital as an alternative to AD (49–51). In our study, using AD as a reference method, data produced using Etest had sensitivity closer to 95%, making it a viable method for fosfomycin testing in addition to disk diffusion. Etests provide MIC values, which may be helpful in clinical decisions if the recently developed novel Etest formulations perform consistently in other laboratories’ hands (51). In that case, CLSI should support studies validating the use of Etest as a susceptibility testing option for fosfomycin, given the numerous challenges of using AD in susceptibility testing and the limitations of disk diffusion.

In conclusion, *in vitro* data from our patient population demonstrate that in addition to *E. coli*, fosfomycin has high activity against several of the most common Gram-negative and Gram-positive bUTI pathogens, including those that are MDR. Furthermore, we’ve shown that the Etest is a viable alternative for fosfomycin susceptibility testing. However, RCTs investigating fosfomycin for the treatment of bUTIs due to organisms other than *E. coli* are lacking whereas clinical, microbiologic, and *in vitro* data strongly suggest this is a viable treatment option. Since bUTIs are among the most common bacterial infections worldwide, occurring in both community and in healthcare settings, and most are usually treated empirically (34), having an effective treatment option against MDR pathogens will play an essential role in managing bUTIs worldwide and antimicrobial stewardship programs. RCTs investigating the use of fosfomycin in treating non-*E. coli* bUTI pathogens and as a combination therapy for treating UTIs, especially in patients with recurrent infections, are long overdue.

## Data Availability

All data produced in the present study are available upon reasonable request to the authors

## ACKNOWLEDGMENTS

Disclaimer: The views expressed in this manuscript are those of the authors and do not necessarily reflect the official policy of the Department of Defense, Department of Army, US Army Medical Department, Defense Health Agency, or the US Government. The study protocol was approved by the appropriate Human subjects regulatory authority. Investigators adhered to the policies for protection of human subjects as prescribed in 45 Code of Federal Regulation 46.

## Ethical statement

The institution’s ethical review board and the human research protection office at the Defense Health Agency and Tripler Army Medical Center reviewed and approved this study.

---

**Supplementary figure 1.**
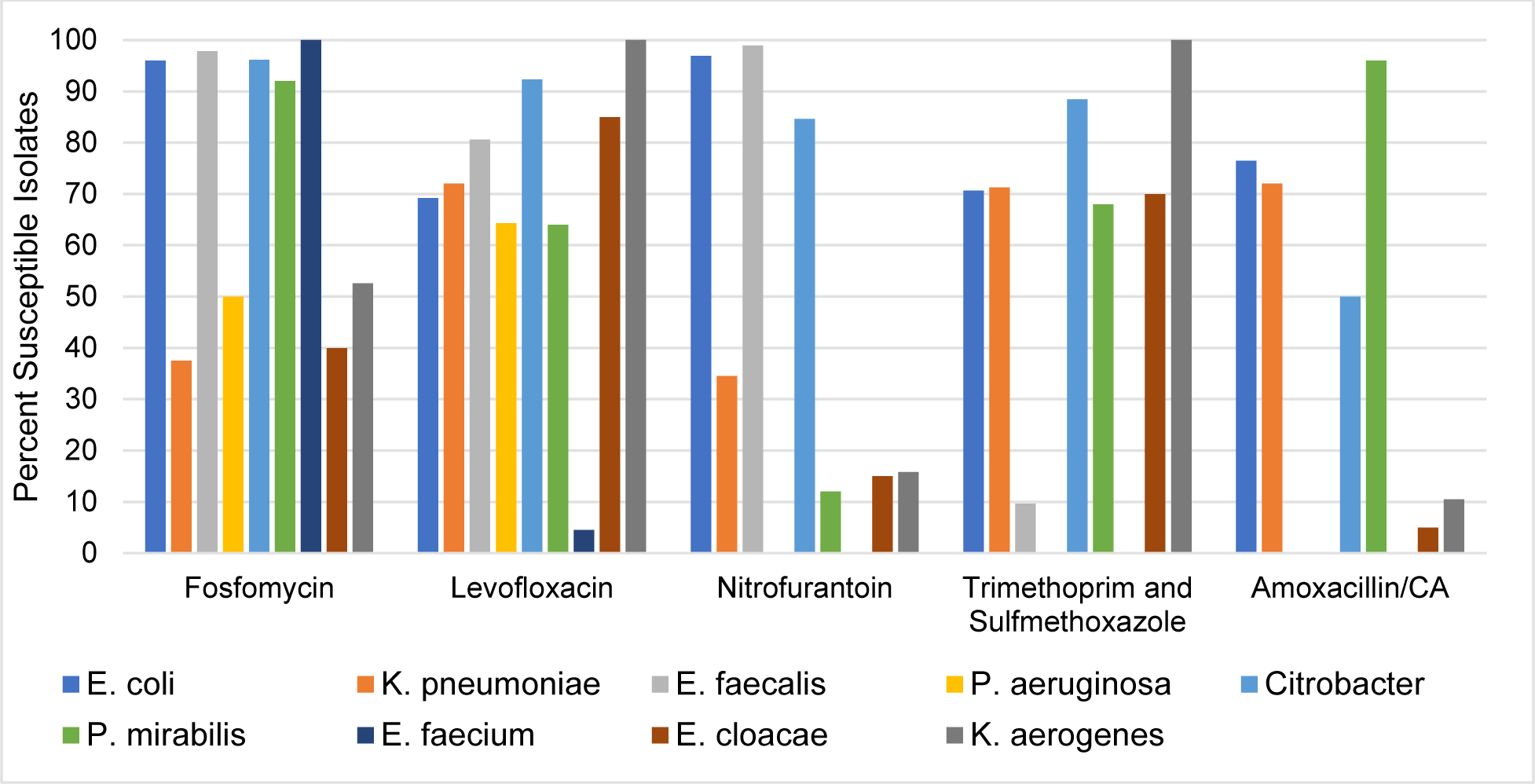
Susceptibilities of clinical isolates to fosfomycin and comparator antibiotics. The agar dilution method was used to test whether clinical isolates were susceptible to fosfomycin. Comparator antibiotic susceptibilities were tested using broth dilution. UTI isolates of *E. coli* (n=890), *K. pneumoniae* (n= 136), and *E. faecalis* (n=93), *P. aeruginosa* (n=28), *Citrobacter* (n=26), *P. mirabilis* (n=25), *E. faecium* (n=22), *E. cloacae* (n=20), *K. aerogenes* (n=19) were tested for susceptibilities to fosfomycin and comparator antibiotics. *E. faecalis* was not tested for amoxicillin susceptibility and *E. faecium* was not tested for nitrofurantoin, TMS, or amoxicillin susceptibilities.

